# A rapid review of the effectiveness and cost effectiveness of interventions that make homes warmer and cheaper to heat for households in fuel poverty in rural and remote areas

**DOI:** 10.1101/2023.04.18.23288747

**Authors:** Deborah Edwards, Judit Csontos, Liz Gillen, Judith Carrier, Ruth Lewis, Alison Cooper, Adrian Edwards

## Abstract

The cost-of-living across the UK has been on the increase since the start of 2021. Living in a rural community is often associated with additional costs compared to those in urban areas. For example, people living in rural areas are not always connected to the gas grid, often using oil and liquid petroleum gas for heating that are more expensive and not subjected to energy price caps. Moreover, housing in rural areas is generally older, not as well insulated and less energy efficient than houses in urban locations, leading to increased risk of fuel poverty. Home energy advice, energy efficiency measures, and financial support all have the potential to mitigate fuel poverty.

The aim of this rapid evidence review was to determine the effectiveness of interventions that make homes warmer and cheaper to heat for households in fuel poverty in rural and remote areas.

Fourteen studies and eight sources of grey literature were included in the review. The included studies and grey literature were published between 2007 and 2022.

There was some evidence of effectiveness for interventions such as energy efficiency home improvements / retrofitting, home improvements (including replacing lightbulbs and electric heaters, external insulation, heating systems, loft insulation installing central heating), Welsh Government Arbed (home improvement and energy efficiency measures) and Nest (home improvements and advice) interventions, provision of energy and home energy advice, and referral for support and/or insulation measures or home improvements. Interventions such as social energy subsidies might not be effective in reducing fuel poverty, and energy efficient social housing was a more efficient method of alleviating fuel poverty. However, the certainty of the evidence is very low, primarily due to the study design and poor quality of the included studies.

Policy makers and funding bodies need to make further investments into research focusing on measures to alleviate fuel poverty, with particular focus on economic analysis. There is a need for high quality, well-developed randomised controlled trials to investigate the effectiveness and cost-effectiveness of energy efficiency measures and advice. Future research should investigate which interventions are the most effective in what types of housing in rural areas to help the targeting of interventions better.

**Funding statement:** The Wales Centre For Evidence Based Care was funded for this work by the Health and Care Research Wales Evidence Centre, itself funded by Health and Care Research Wales on behalf of Welsh Government.

## 1. BACKGROUND

### 1.1 Who is this review for?

This Rapid Review was conducted as part of the Health and Care Research Wales Evidence Centre Work Programme. The above question was suggested by Technical Advisory Cell (TAC), Welsh Government, and Taf Housing.

### 1.2 Background and purpose of this review

The cost-of-living across the UK has been on the increase since the start of 2021, reflecting the position globally. Road fuel prices and household energy bills have risen due to the effect of Russia’s invasion of Ukraine (Francis-Devine et al. 2022).

Fuel poverty is defined as “spending more than 10% of a household’s income on fuel for satisfactory heating and comfort and to sustain all energy services” (Charlier & Legendre 2021, p.121557). The main drivers of fuel poverty are energy prices, energy efficiency, low household income and how energy is used in the home (Scottish Government 2016). The consequences of fuel poverty and low incomes identified prior to the current cost of living crisis, for example living in a cold damp home, have a negative impact on both physical and mental wellbeing (Chakravorty 2022, Volkos & Symvoulakis 2021, Thomson et al. 2022, Roberts et al. 2022). It is envisaged that the number of excess winter deaths will increase as fuel poverty rates increase (Roberts et al. 2022).

In Wales, although the current cost-of living crisis effects all households, living in a rural community is often associated with additional daily living costs compared to those living outside of rural areas (Senedd Cymru 2022a, Roberts et al. 2022). In 2020, many domestic properties were not connected to the gas grid, the highest percentage being in rural areas including mid and west Wales (DBEIS 2021), it has been reported that nearly a third of rural households use oil as their main fuel for heating (Wales Audit Office 2019). Compared to networked energy sources, oil and liquid petroleum gas are more expensive and are not subjected to energy price caps (Senedd Cymru 2022a). A further concern is that rural housing is generally older, not as well insulated and less energy efficient than houses in urban locations (Senedd Cymru 2022a, Senedd Cymru 2022b). Higher petrol and diesel costs in rural filling stations, alongside a lack of available public transport leading to higher use of privately owned motor vehicles are also all contributing factors to rural fuel poverty (Senedd Cymru 2022a).

Programmes to improve how energy is used in the home (home energy advice), to improve the energy performance of homes, financial support and community level energy buying clubs and cooperative arrangements all have the potential to mitigate fuel poverty (Javornik & Mackie 2022, Powell et al. 2018) which in turn can result in health improvements and improved quality of life (Powell et al. 2018). Interventions can be large scale government subsidised activities focusing on improving the energy efficiency of housing stock and or household appliances or locally delivered projects through partnerships with local authorities, housing providers or third sector organisations. The aim of this rapid review was to determine the effectiveness of interventions that make homes warmer and cheaper to heat for households in fuel poverty in rural and remote areas.

## 2. RESULTS

### 2.1 Overview of the Evidence Base

The evidence base consists of primary research evaluations presented across 14 studies which consisted of one randomised control trial (with partial crossover) (Heyman et al. 2011), three quasi-experimental studies (Grey et al. 2017, Poortinga et al. 2017, Willand et al. 2019); two pre-test / post-test with a control group (Shortt & Rugraska 2007, Wade et al. 2019); two pre-test / post-test with no control group (Eadson & Leather 2017, Papada et al. 2021); one post-test with control group (Sharpe et al. 2020), one case study (McGinley et al. 2022); one case control study (Charlier et al. 2019) and three quantitative descriptive surveys (Miller et al. 2022, Sherriff et al. 2020, Welsh Government 2015). Five of these were part of wider mixed methods studies (Shortt & Rugkåsa 2007, Miller et al. 2022, Wade et al. 2019, Welsh Government 2015, Willand et al. 2019). Additional information (n=8) has been provided by UK grey literature sources including the evaluations and/or annual reports of the Welsh Government Warm Homes Program - Nest (Welsh Government 2022) and Arbed (Arbed am Byth 2022, Patterson 2012), Scottish schemes (Citizens Advice Scotland 2016, Citizens Advice Scotland 2017, Shelter Scotland 2017), UK schemes (DECC 2015) and English schemes (NEA 2019). Of these, four are catalogues of fuel poverty schemes (Citizens Advice Scotland 2016, DECC 2015, NEA 2019, Shelter Scotland 2017).

Only five of the evaluations focused entirely on rural properties and these were conducted in Scotland (Sherriff et al. 2020, Wade et al. 2019); Ireland (McGinley et al. 2022); Northern Ireland (Shortt & Rugkasa 2007) and Cornwall, England (Sharpe et al. 2020). A further evaluation was conducted in a remote region of Greece (Papada et al. 2021). The remainder of the evaluations (n=8) were conducted across both urban and rural areas throughout Wales (Grey et al. 2017, Miller et al. 2022, Pootinga et al. 2017; Welsh Government 2015) and England (Eadson & Leather 2017, Heyman et al. 2011); France (Charlier et al. 2019) and Australia (Willand et al. 2019). Most grey literature (n=7) covered both urban and rural areas across the UK, except the report by Shelter Scotland (2017) which focused mainly on rural areas.

The interventions that were evaluated were home energy advice, referral for support and/or insulation measures or home improvements (including Nest) (Eadson & Leather 2017, Sherriff et al. 2020, Wade et al. 2019, Welsh Government 2015), social energy subsidies and energy efficient housing (social housing) (Charlier et al. 2019); energy efficiency home improvements / retrofitting (Heyman et al. 2011, McGinley et al. 2022, Willand et al. 2019) (including Arbed)(Grey et al. 2017, Poortinga et al. 2017, Miller et al. 2022); central heating (Sharpe et al. 2020, Shortt & Rugkasa 2007); energy related living lab - advice with monitoring equipment (Papada et al. 2021). The grey literature evaluations and/or annual reports focused on energy efficiency home improvements and advice (n=1) (Welsh Government 2022), on energy efficiency home improvements / retrofitting (n=2) (Arbed am Byth 2022, Patterson 2012), advice (Citizens Advice Scotland 2017). The catalogues all described a variety of energy efficiency measures and other fuel poverty interventions (Citizens Advice Scotland 2016; DECC 2015, NEA 2019; Shelter Scotland 2017).

### 2.2 Effectiveness of interventions

The effectiveness of **social energy subsidies** (energy subsidy voucher of €48 to €227 a year, depending on the household income and composition) and **energy efficient housing** (social housing) to reduce fuel poverty was assessed by Charlier et al. (2019). The sample sizes of those in receipt of the social energy subsidy was small (n=81) compared to those in receipt social housing (n=422) and those in the control group (n=552), statistical analyses were poorly presented, there were inconsistencies in the descriptions of treatment and control groups. However, it was determined that providing social energy subsidies did not alleviate fuel poverty. Providing energy efficient housing (social housing) was found to be a more efficient means of alleviating fuel poverty.

Energy-related living lab which aims to provide low-cost methods to energy vulnerable households in order to tackle fuel poverty was the focus of the study by Papada et al. (2021). The living lab consisted of **installing monitoring equipment in the homes**, **inspecting heating systems and providing energy specific advice**. Households that had monitoring equipment installed, energy advisor visits and advice were significantly more likely to apply energy efficiency measures (p=0.001) compared to those who did not have energy monitoring equipment installed. Households also reported a positive change in their energy related behaviours and an improvement in their quality of life (improved thermal comfort, facing less moisture problems and reduced energy costs) compared to those who did not have energy monitoring equipment installed. Additionally, due to the maintenance of central heating systems in 12 of the households a significant increase in the burners’ energy-efficiency ratio was detected, resulting in considerable energy savings and a reduction of the households’ energy costs.

Heyman et al. (2011) conducted a randomised controlled trial (with partial crossover) to assess the personal, social and economic benefits of **energy efficiency measures** in the form of **housing improvements** (loft insulation, cavity wall insulation, draught exclusion, heating controls and central heating). Households that had received energy efficiency measures demonstrated improved Standard Assessment Procedure (SAP) ratings by 12 points compared to the control group (p<0.001). This translated into the households with energy efficiency measures generating increases in winter evenings living room temperatures of one to two degree Celsius compared to the control group (p=0.03). The greatest temperature increases were associated with the combination of heating system and insulation measures. Families did not respond to energy efficiency gains by reducing their heating expenditure. Because homes receiving energy efficiency measures were more fuel-efficient than those of the control group, the former could spend less on fuel than the latter in order to achieve the same room temperature. However, households with energy efficiency measures seemed to have increase temperatures, choosing increased warmth over reduced bills.

A post-intervention survey was used to assess the impact of an intervention to install a new first time **central heating system** in order to reduce fuel poverty (Sharpe et al. 2020). Responses were compared between a waiting list control and those who had central heating installed for the first time. Significantly (p<0.01) more households (50.75%) in the intervention group reported that there had been an improvement in their ability to pay their bills compared to the control (28.21%) Additionally, significantly (p<0.01) less households (58.75%) in the intervention group reported that they had to avoid heating their home due to costs compared to the control (83.54%).

A study conducted in rural areas in Northern Ireland (Shortt & Rugkasa 2007) evaluated the **installation of central heating** for households in fuel poverty. It was reported that there was relatively little change in average indoor temperature after installing central heating although this was only measured across 12 of the 54 households that were in receipt of heating installation and not compared with the control households. Average fuel costs of households in the intervention group significantly decreased from £1113 per year to £751.56 (p<0.001). Another objective of the study was to encourage people who were eligible for benefits, but not claiming them, to do so. However, this was inaccurately and poorly reported and no conclusions can be drawn.

The retrofitting of five detached properties in rural Ireland was investigated by McGinley et al. (2022). The authors reported that due to the differences in case study properties and measures installed **(external insulation, heating systems, loft insulation)** that it was difficult to make comparisons across the properties. However, each property experienced various benefits from **retrofitting**, including increased thermal comfort (ranging from 1% (case D) to 19% (case A) increase in indoor temperature), and reduced energy usage (ranging from 1% (case C) to 66% (case B) reduction in primary energy usage).

Willand et al. (2019) conducted a quasi-experimental study as part of a wider mixed methods investigation to quantify changes in indoor temperatures, energy consumption, energy costs and health due to building retrofits. The findings showed that the households who had received **energy efficiency retrofit improvements** intervention had improved their mean energy efficiency star rating from 0.8 stars to 3.5 stars. However, data was only available for a sub-sample of properties and no statistical analysis was performed to compare this to households in the control group. Valid indoor temperature data was available for 12 living rooms and 12 bedrooms. Taking only occupied days of the home into account, the retrofit intervention significantly reduced electricity consumption (p=0.17) but not gas consumption (p=0.742). These benefits were primarily attributed to the replacement of light bulbs with light-emitting diode (LED) lights and of portable electric heaters with new reverse cycle air conditioners (RC ACs). However, sample sizes were too small, as statistical analysis was only performed on sub samples of households (n=10 or less), and as such are underpowered.

#### 2.2.1 Welsh Government Warm Homes Program (Arbed and Nest)

The Welsh Government [Warm Homes Programme](https://www.gov.wales/get-help-energy-efficiency-your-home-nest) funds energy efficiency improvements to those households who are eligible alongside free advice to all households in Wales. Improvements are currently delivered through the Nest scheme and previously through the Arbed Scheme.

The Arbed scheme started in 2009 until November 2021 (Audit Wales 2021) and was an area-based scheme which offered energy efficiency improvements in targeted areas (Welsh Parliament 2022). Two quasi-experimental studies investigated the impact of the Arbed scheme on a number of outcomes and including thermal satisfaction, fuel poverty, financial difficulties, financial stress (Grey et al. 2017) and internal conditions and household energy use (Poortinga et al. 2017). Grey et al. (2017 reported that respondents who received **energy efficiency measures** to their homes through the scheme reported fewer financial difficulties, higher thermal satisfaction, and lower levels of fuel poverty meaning they were less likely to put up with the cold to save money on heating. However, the authors only selectively reported key statistical significant findings and did not report on financial stress. Poortinga et al. (2017) conditions monitored internal conditions for a minimum of 28 consecutive days before and after the installation of **energy efficiency measures** and this was then compared with households that were not in receipt of Arbed. It was demonstrated that households in receipt of Arbed had a significantly increased indoor air temperature by on average 0.84 K compared to the households in the control areas (p<0.05). This resulted in the bringing the majority of indoor temperature measurements within the ‘healthy’ comfort zone of 18–24°C and a drop in average daily gas usage of 37%. The different intervention measures had varying effects on indoor room temperature, with external wall insulation being the most effective measure. Additionally, the greatest increases were found in the evening and at night, in the bedroom, and in British steel-framed buildings.

The annual report (Patterson 2012) for the of the first scheme (Arbed 1) postulated that households could save on their energy bills as a result of the installation (assuming that their energy behaviour remained the same post installation). With regard to improvements in the energy performance rating, the average SAP before the installation across all the properties was 60 (range 43 to 66) compared to 69 following the works (range 58 to 82). This improvement was also reflected in the average annual energy bill which was £990 before works were undertaken compared with £774 after. A cross sectional survey of the evaluation of Arbed 3 reported similar findings (Miller et al. 2022). Householders self-reported a reduction in energy costs post-installation, demonstrating the likely impact the measures had on increasing monthly disposable income. Following installation, 61% of householders claimed that they now spent less than 10% of their income on energy bills (compared with 31% pre installation) taking these households out of fuel poverty. This cost saving was also reflected in those households categorising themselves as being in severe fuel poverty (spending more than 20% of their income on energy bills) a self-reported reduction from 30% pre-installation to 5% post-installation. The report summarised that it is these households that appeared to benefit from the greatest proportional savings. Additionally, all households where funded measures were installed experienced an EPC uplift of at least one band and SAP ratings were also increased.

The final Arbed annual report (Arbed am Byth 2022) before the scheme closed reported the following:

- 5,050 measures were installed in homes,
- 1,032 properties were treated,
- 2,395 whole house assessments were carried out in order to complete the Energy Performance Certificate (EPC),
- the average improvement in the energy performance rating (SAP) of each property was 18.93 points,
- customer satisfaction was rated at 100%.

The Nest scheme started in 2011 (Audit Wales 2021) and is a need-based scheme for fuel poor households who are in receipt of a means tested benefit and who live in a very energy inefficient home, with a SAP rating of F or G. The scheme offers package of free home energy improvements as well as advice, on saving energy, money management, fuel tariffs, benefit entitlement checks and referral to alternative schemes to all householders in Wales (Welsh Parliament 2022).

In 2014, a mixed methods independent evaluation of the scheme was commissioned (Welsh Government 2015). A total of 18,481 measures were installed up to September 2014, in 15,603 households. Gas boilers accounted for the majority of measures (around two-thirds of all interventions), followed by oil (11%) and loft insulation (10%). However, the geographical reach of the scheme was reported to be uneven and the targeting of rural areas was a challenge. Householders felt better able to heat their homes as a result of being given **advice** (35%) or receiving home improvement measures (89%). Participating households receiving **home improvements** reported a reduction in energy bills (62%) and were aware of their energy use (83%). In terms of value for money it was estimated that the overall annual energy saving across all the households that took part in the scheme up until the time of the evaluation had been £7.48m.

From the most recent annual report (2020-21) it was estimated that Nest home energy efficiency improvements have delivered energy bill savings averaging £305 per household per year (Welsh Government 2022). Other benefits of interest included:

- benefit entitlement checks resulted in households that are now eligible for new or additional benefits (average £2,091 potential increase in benefit take-up per household),
- 3,458 customers were referred to their energy supplier for Warm Homes Discount and of these 366 Nest customers qualifying for the discount, amounting to total savings of £51,240,
- 2,025 customers received money management advice and additionally 1,274 received debt advice,
- 99% of customers reported satisfaction with advice and installations provided by Nest.

#### 2.2.2 UK schemes

In 2015, the Department of Energy and Climate Change (DECC) commissioned the National Energy Action (NEA) to carry out an online survey to catalogue local schemes that are targeting individuals with health problems for energy efficiency measures and other fuel poverty interventions (DECC 2015). Seventy five unique schemes across England and Wales were identified and follow up interviews were conducted with 19 (out of 21). The geographical scope of the schemes was either focused on local authority, regional or national areas with only seven schemes were conducted in rural locations and just under half in rural and urban locations. The schemes included the following services: low cost energy efficiency measures, medium to high-cost energy efficiency measures, energy-related advice, referral to energy-related grants, support and advice and referral to other services. The catalogue provided information on whether the schemes had been evaluated (47%: 36/76) but no further detail was provided. The outcomes measured and reported against across these schemes were the ability to heat the home, including the proportion of income spent on fuel, applying for benefits, trust fund grants secured, energy debt cleared, and energy savings made (£’s and kilowatts per hour).

Citizen Advice Scotland catalogued and reviewed past and current energy efficiency and fuel poverty schemes in the UK (Citizens Advice Scotland 2016). The catalogue included UK-wide supplier obligations (3 schemes); other UK-wide energy efficiency schemes (2 schemes); UK-wide cash-benefits schemes (3 schemes); and renewable energy schemes (6 schemes). Only brief details of any evaluation were provided for three of these schemes.

#### 2.2.3 English schemes

The NEA updated the DECC 2015 catalogue in 2019 and presented information for 34 health-related fuel poverty schemes in England and noted that although there appear to be widespread activity that interventions were patchy and represented a “post code lottery” (NEA 2019). No detail was provided regarding any evaluation of the schemes. The approach to monitoring and evaluation was noted as part of this review.

The impact of the Fuel Poverty and Health Booster Fund on keeping homes warmer and keeping up with energy bills was evaluated by Eadson & Leather (2017). This sought to provide home energy advice and referral for support and/or insulation measures to households in fuel poverty living across nine local authorities in England. Questionnaire responses from pre and post survey indicated that the majority of participating households (63%) reported finding it very easy to keep their homes warm post intervention compared to 5% pre intervention. Regarding keeping up with energy bills, more participants reported to be able to manage well (25%) or quite well (38%) post intervention, compared to pre-intervention when only 6% and 20% reported managing. However, no statistical analysis was completed, thus it is unclear whether changes detected were significant.

#### 2.2.4 Scottish schemes

In September 2016, Shelter Scotland and Energy Action Scotland partnered to create a catalogue of health-related fuel poverty schemes which followed a similar approach to the NEA 2015 catalogue (Shelter Scotland 2017). Thirty-one schemes were identified, and it was noted that evaluations had been conducted and or reported for all apart from five but no further detail was provided (Shelter Scotland 2017). Additionally, Citizens Advice Scotland (2016) catalogued 12 Scotland-specific energy efficiency and fuel poverty schemes as part of their review of energy efficiency and fuel poverty schemes in the UK. However, no formal evaluation was conducted for these schemes. The authors did conclude that although increased energy efficiency helps to mitigate fuel poverty, that it is not sufficient on its own to eliminate it, with many low-income consumers in more efficient houses still remaining in fuel poverty.

An overview of 158 face-to-face fuel poverty projects across Scotland that provided advice to households was provided by Citizens Advice Scotland (2017). The advice that was offered included fuel debt, tariffs and suppliers, energy behaviours, heating systems, other billing issues and referrals for energy efficiency measures. Although this was a mixed methods evaluation the findings were mainly qualitative. However, billing savings achieved per client were reported across 30 projects with average bill savings of £316.

Wade et al. (2019) evaluated the Home Energy Scotland (HES) Homecare pilot programme, which focused on the Energycarer approach to tackle fuel poverty in rural Scotland. Energycarers acted as case workers providing individually tailored solutions for households in fuel poverty based on home and needs assessment, looking at heating or insulation measures that could be installed, and householders ability to pay for improvements or whether they qualify for the Warmer Homes Scotland programme. Energycarers usually visited households three to four times, but more appointments could be arranged based on participants’ needs. To evaluate this pilot programme, a pre-test / post-test study with a control group was conducted, focusing on internal temperature and thermal comfort changes. Internal temperature did not change significantly following installation of individually tailored measures (gas boiler, draughtproofing, electric storage heater) based on the Energycarers interventions. However, as the sample size is too small (n=3), this statistical comparison is not appropriate. Thermal comfort based on a questionnaire was descriptively analysed, and while some improvements were noticed following the Energycarer intervention, due to insufficient sample, variation in households and between intervention and control groups no conclusions can be drawn from the evaluation.

The Gluasad Còmhla (Moving Together) project, which aimed to provide energy advice and assistance with energy efficiency home improvements as social prescribing in the Outer Hebrides, Scotland was evaluated by Sherriff et al. (2020). A descriptive survey was conducted as part of a wider mixed methods study, and the questionnaire focused on householders’ use of the heating system, self-reported internal temperature, cost of running the heating system, money available after bills, and the indoor temperatures effect on people’s activity levels. Findings suggest that majority of respondents used the heating system about the same (37%) or slightly less often (22%) following the intervention compared to pre-intervention. However, they felt their home was much (26%) or slightly warmer (30%), indicating improvements. Most respondents (33%) reported paying about the same or slightly less (26%) following the Moving together intervention, whilst reporting that they had about the same amount of money after paying bills (30%) or slightly more (22%). Responding to the question about the effect of the indoor temperature on activities, over half of the respondents (56%) said it affected them about the same as prior to the intervention. However, the sample size was small and data was only collected after the intervention, so it cannot be determined whether the intervention had a significant impact on making homes warmer and cheaper to heat.

#### 2.2.5 Bottom line results for effectiveness of interventions

This section summarised the effectiveness of interventions, including subsidies, energy efficiency home improvements / retrofitting, advice, and a living lab from 14 studies and from from eight evaluations and/or annual reports. The overall certainty in the evidence was assessed based on seven studies that provided robust methods and statistical analysis. All other evidence was classed as ungraded.

- Ungraded evidence suggests that interventions involving home energy **advice, referral for support and/or insulation measures or home improvements may make homes warmer, cheaper to heat and enable householders to keep up with energy bill payments.**
- Ungraded evidence suggested that **social energy subsidies** might not be **effective in reducing fuel poverty**, and that **energy efficient social housing** was a **more efficient method** of alleviating fuel poverty.
- Very low quality evidence demonstrated that installing **central heating significantly improved** householders’ **ability to pay energy bills**, **reduced energy costs** and **significantly less households avoided heating their homes** due to costs, although **very little change was reported in average indoor room temperature**.
- Very low quality evidence showed that householders receiving **energy efficiency home improvements** as part of the Welsh Government Warm Home Scheme - **Arbed** experienced significantly higher indoor air temperatures, increased indoor temperatures reaching the healthy comfort zone (18-24^0c^), **improved thermal satisfaction**, **reductions in average daily gas consumption, fewer financial difficulties**, and **fewer respondents reported putting up with feeling the cold to save heating** compared to the control group. Further ungraded evidence suggests that householders who were in receipt of Arbed experienced a **reduction in their energy costs,** and improved household energy efficiency ratings **(increased their EPC band and SAP rating).**
- Ungraded evidence suggested that the Welsh Government Warm Home Scheme – **Nest** which delivered **home improvements and energy advice** enabled householders to **better heat their homes**, **reduced their energy bills** and **increased their awareness of their energy use.**
- Very low quality evidence showed that **energy efficiency home improvements / retrofitting** led to significantly **improved household energy efficiency ratings (SAP ratings)**, and **increased living room temperatures**, although **energy bills did not reduce** significantly compared to the control group.
- Very low quality evidence demonstrated that home improvements, such as **replacing lightbulbs and electric heaters increased energy efficiency ratings** (star ratings) and **reduced electricity consumption**, but **no change** was detected **in gas consumption**.
- Ungraded evidence **suggested that** home improvements increased thermal comfort and reduced energy consumption.
- Ungraded evidence suggested that providing **energy related advice** can make an impact on **energy bill savings.**
- Very low quality evidence showed that providing monitoring equipment in the homes, inspecting heating systems and **providing energy specific advice** (a living lab intervention) **significantly increased the likelihood** of householders’ installing **energy efficiency measures.** Additionally, **improved energy savings, reduced costs** and **energy related behaviour changes were reported.**

## 3. DISCUSSION

### 3.1 Summary of the findings

The interplay of four factors including low income, high energy costs, insufficient energy performance, and high energy usage in the home environment can lead to fuel poverty (Scottish Government 2016). Thus, improving energy performance of houses is crucial in fighting fuel poverty, and government initiatives focusing on energy efficiency both in the UK and abroad have had some success. The findings of this rapid review indicate that **energy efficiency interventions,** such as external insulation, central heating installation, and heating system changes, can have a beneficial impact on householders’ ability to heat their homes and reduce energy costs. Furthermore, energy efficient social housing was found more effective in alleviating fuel poverty than social energy subsidies (Charlier et al. 2019). However, increases in fuel prices and insufficient income raises can influence the amount of benefit gained from interventions aiming to improve fuel poverty (Scottish Government 2016).

A number of schemes as reported in the catalogues across the UK incorporate providing energy advice to those in fuel poverty regarding topics, such as fuel debt, tariff and suppliers, energy behaviours, heating systems, and they can make referrals for energy efficiency measures schemes (Citizens Advice Scotland 2016, DECC 2015, NEA 2019, Shelter Scotland 2017). Where the outcomes of evaluations have been reported, households who act on the advice given are better able to heat their homes and report a reduction in average bill savings (Papada et al. 2021, Welsh Government 2015, Welsh Government 2022, Citizens Advice Scotland 2017).

This rapid review aimed to look for interventions in rural areas, although only three of the 11 included evaluations addressed rural fuel poverty specifically. Powell et al. (2018) in reporting an evidence review of interventions to address rural fuel poverty across OECD countries also identified very little specific evidence for rural areas and only tentative conclusions regarding the effectiveness of the interventions could be made. However, this mainly included grey literature reports with limited peer-reviewed evidence, in contrast with this rapid review.

Previous reviews have mainly described schemes that have the potential to address fuel poverty (Das et al. 2022) or that have investigated different types of interventions that encourage energy conservation across all households and not just those in fuel poverty (Das et al. 2022, Delmas et al. 2013, McAndrew et al. 2021). This rapid review is unique as it specifically looked at studies that investigated the impact of interventions on fuel poor populations.

### 3.2 Strengths and limitations of the available evidence

The included studies had several limitations based on the methodological assessment. Of the 14 included studies, one randomised controlled trial (Heyman et al. 2011) scored six out of a potential ten criteria on the critical appraisal checklist. Three questions were not applicable as the blinding of participants (Q4), or those delivering the intervention or assessing the outcome (Q5, Q6) was not feasible. The study was down scored as it was unclear if true randomisation was used in the assignment of participants (Q1) or if the allocation to treatment groups was concealed (Q2). Moreover, treatment groups were not similar at baseline (Q3).

Seven of the included studies were appraised by using the JBI checklist for quasi-experimental studies. Two of these, (Grey et al. 2017, Poortinga et al. 2018) met eight out of the potential nine critical appraisal criteria. However, the remaining four reports scored four (Eadson & Leather 2017, Papada et al. 2021, Shortt & Rugkasa 2007) and five (Wade et al. 2019; Willand et al. 2019). It was unclear if the participants included in any comparisons received similar treatment other than the intervention of interest (Q3). One of the studies (Papada et al. 2021) did not include multiple measurements of the outcome pre and post intervention (Q5). The other two studies either did not report or it was unclear whether outcomes were measured in the same way (Q7). Shortt & Rugkasa (2007) also had issues with the reliability of the measures used (Q8) and Wade et al. (2019) conducted statistical analysis on a sub sample of three households. Moreover, Eadson & Leather (2017) did not clarify whether the same participants or two different samples were compared descriptively, and no explanation was given regarding why inferential statistics were not conducted.

One study appraised with the JBI checklist for cohort studies (prospective) (Sharpe et al. 2020) scored only four out of a potential nine criteria. Although the groups were similar upon recruitment (Q1) and exposure was measured similarly to assign participants to groups (Q2) and in a valid and reliable way (Q3), any confounding factors or strategies to deal with confounding were not identified or stated (Q4, Q5). Furthermore, it was unclear if the participants were free of the outcomes at the start of the study (Q6) or if outcomes were measured in a valid and reliable way (Q7).

Of the remaining five included studies, one case report (McGinley et al. 2022) scored highly, meeting seven out of the eight critical appraisal criteria. Three studies were appraised using the JBI checklist for analytical cross-sectional studies (Sherriff et al. 2020, Miller et al. 2022, Welsh Government 2015) and they all score five out of a potential eight criteria. They did not report identifying confounding factors and strategies to deal with them (Q5, Q6). Finally, one case control study (Charlier et al. 2019) met five of the potential 10 criteria as it was unclear whether groups were comparable at the start of the study (Q1), and if the exposure was measured reliably and in the same way for all (Q5, Q6).

As evidenced, many of the included studies had quality issues in terms of being comparable at entry and any interventions and outcomes being measured reliably and consistently across groups. There was a lot of variation within the reports, in terms of the type of housing included in the sample (Papada et al. 2021), period of construction (Charlier et al. 2019), along with any measures installed, outcomes assessed, such as reduction in energy consumption, change in room temperature or thermal comfort. This made it difficult for studies to compare results across properties. It has been noted that this is also an issue across the wider evidence base in this field (Delmas et al. 2013, McAndrew et al. 2021). Additionally, occupancy and occupant behaviour, which can have a large impact on energy consumption were often not considered (Poortinga et al. 2017). Furthermore, the geographical scope of the studies reviewed varied, focusing on local authority, regional or national areas. At times this, together with the general lack of reporting, made it difficult to categorise studies as rural or urban.

Some studies presented no statistical analysis whilst others poorly presented the information with two studies only performing statistical analysis on a sub sample of households (Wade et al. 2019; Willand et al. 2019) and another only selectively reporting the key statistically significant findings (Grey et al. 2017). Sample size was an additional limiting factor for many of the included reports, with small samples restricting the reliability of the findings and the ability to draw conclusions regarding the effectiveness of the interventions for larger population groups. There was also an absence of any sample size calculations, highlighting a further need for caution when interpreting report findings.

There is a notable absence of economic evaluations of housing improvements within the included studies. This was also noted by Fenwick et al. (2013) who conducted a systematic review of economic analyses on the health impacts of housing improvements, and the majority of included studies (n=25/29) presented data on intervention and/or recipient costs only and, despite sufficient data, opportunities to conduct economic analysis have been missed. Furthermore, Thomson et al. (2009) mentioned in their systematic review whether included studies had reported some economic analysis. While some socioeconomic benefits were mentioned, Thomson et al. (2009) concluded that economic impact assessments should be planned alongside studies looking at the effect of housing improvements.

### 3.3 Implications for policy and practice

There is a need for high quality research, particularly for well-developed randomised controlled trials that investigate the effectiveness and cost-effectiveness of measures, such as external insulation, heating system changes, with careful consideration to confounding factors. The interventions will likely need to be multicomponent. However, identifying individual measures that are most effective, or redundant, is also important, which may require innovative adaptive trial design or analysis.

Future research should investigate which interventions are the most effective in what types of housing in rural areas to help the targeting of interventions better.

Policy makers and funding bodies need to make further investments into developing funding calls for investigating measures to alleviate fuel poverty, with particular focus on economic analysis.

### 3.4 Strengths and limitations of this Rapid Review

Strength of this rapid review is that systematic, protocol-driven procedures were followed in the searching and identification of key literature, supported by an experienced information specialist. Moreover, websites of relevant UK organisations and governments were searched to identify grey literature reports and evaluations. However, as this is a rapid review, some of the review processes are streamlined and modified to ensure timely production. Approximately Title and abstract screening of the citations retrieved from the database searches were conducted by one reviewer and approximately, 20% for consistency and accuracy by a second reviewer, therefore some potentially relevant studies might have been missed. All full text screening was conducted by one reviewer and checked by a second reviewer. Critical appraisal of all evidence was conducted by one reviewer and checked by a second reviewer.

As detailed above due to the quality of included studies, heterogeneity of interventions, outcomes and outcome measures used, it is not possible to provide strong implications for policy and practice on which interventions aiming to reduce fuel poverty in rural areas are the most beneficial. While, this review has its limitations, the included evidence consistently shows that energy efficiency measures might be beneficial to keep houses warm and alleviate fuel poverty, indicating potential for these interventions.

## Data Availability

All data produced in the present study are available upon reasonable request to the authors

## Abbreviations

DECC: Department of Energy and Climate Change
DBEIS: Department for Business, Energy and Industrial Strategy
EPC: Energy Performance Certificate
HES: Home Energy Scotland
LED: Light-emitting diode
NEA: National Energy Action
OECD: Organisation for Economic Co-operation and Development
RC Acs: Reverse cycle air conditioners
SAP: Standard Assessment Procedure
TAC: Technical Advisory Cell

## 5. RAPID REVIEW METHODS

### 5.1 Eligibility criteria

Inclusion criteria were informed by the PICOS (Population, Intervention, Comparison, Outcome, Study design) framework. Inclusion criteria were also limited to high income countries (HICs), as research findings from low- and middle-income countries (LMICs) might not be fully transferable to the UK context. To check countries income status, the World Population Review website (https://worldpopulationreview.com/country-rankings/high-income-countries) was used.

**Table 1:**
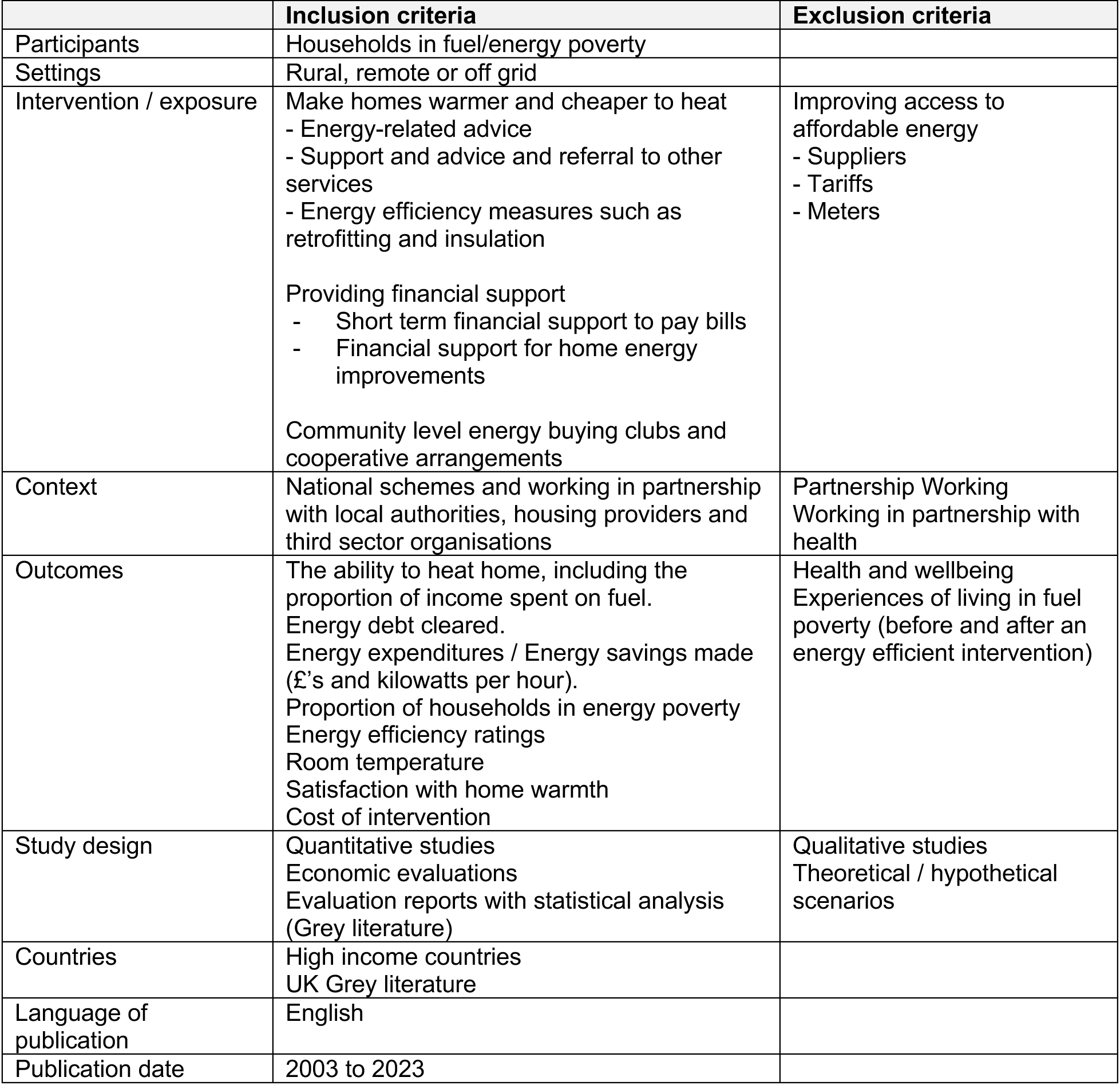
Eligibility criteria.

|  | **Inclusion criteria** | **Exclusion criteria** |
| --- | --- | --- |
| Participants | Households in fuel/energy poverty |  |
| Settings | Rural, remote or off grid |  |
| Intervention / exposure | Make homes warmer and cheaper to heat<br>- Energy-related advice<br>- Support and advice and referral to other services<br>- Energy efficiency measures such as retrofitting and insulation<br>Providing financial support<br>- Short term financial support to pay bills<br>- Financial support for home energy improvements<br>Community level energy buying clubs and cooperative arrangements | Improving access to affordable energy<br>- Suppliers<br>- Tariffs<br>- Meters |
| Context | National schemes and working in partnership with local authorities, housing providers and third sector organisations | Partnership Working<br>Working in partnership with health |
| Outcomes | The ability to heat home, including the proportion of income spent on fuel.<br>Energy debt cleared.<br>Energy expenditures / Energy savings made (£'s and kilowatts per hour).<br>Proportion of households in energy poverty<br>Energy efficiency ratings<br>Room temperature<br>Satisfaction with home warmth<br>Cost of intervention | Health and wellbeing<br>Experiences of living in fuel poverty (before and after an energy efficient intervention) |
| Study design | Quantitative studies<br>Economic evaluations<br>Evaluation reports with statistical analysis (Grey literature) | Qualitative studies<br>Theoretical / hypothetical scenarios |
| Countries | High income countries<br>UK Grey literature |  |
| Language of publication | English |  |
| Publication date | 2003 to 2023 |  |

### 5.2 Literature search

#### 5.2.1 Evidence sources

Searches for research material was conducted across four databases: Embase (on the OVID platform), ASSIA, Scopus, and Web of Science, from 2003 to January 2023 for English language citations. Searches for UK grey literature was conducted using relevant organisational websites (see Appendix 1) and further potentially relevant reports can be found in Appendix 2.

#### 5.2.2 Search strategy

An initial search of Scopus had been undertaken as part of the rapid evidence summary (December 2022) that informed the rapid review. The key words used within the title of a publication were fuel poverty OR energy poverty AND efficiency measure OR intervention OR initiative OR program* OR polic* or strategy* OR service* AND rural OR remote. An analysis of the text words contained in the title and abstract, and of the index terms used to describe each article had been then conducted to inform the development of a search strategy which was tailored for each information source. The complete search strategy is presented in the Additional material. The reference list of all included studies was screened for additional papers. Due to time constraints, we deviated from the protocol and we were unable to conduct forward citation searching for all included studies.

#### 5.2.3 Reference management

All citations retrieved from the database searches were imported or entered manually into EndNote^TM^ (Thomson Reuters, CA, USA) and duplicates removed. Irrelevant citations were removed by searching for keywords within the title using the search feature within the Endnote software. The project team agreed which keywords to use to identify papers which did not meet the inclusion criteria. At the end of this process the citations that remained were exported as a text file in Endnote export style and then imported to Rayyan^TM^.

### 5.3 Study selection process

The citations were screened by a single reviewer with keyword categories for include, exclude highlighted using the software package Rayyan^TM^. Two reviewers dual screened at least 20% of citations using the information provided in the title and abstract resolving all conflicts if needed.

For citations that appeared to meet the inclusion criteria, or in cases in which a definite decision could not be made based on the title and/or abstract alone, the full texts of all citations were retrieved. Full-text documents were checked by a single reviewer with a screening tool developed for this rapid review containing questions about the inclusion criteria. The screening tool had been piloted on full-text documents found during initial searches, and changes had been made when necessary to make the screening tool fit for purpose. A second reviewer double checked the full-text documents and made a final decision. The flow of citations through each stage of the review process were displayed in a PRISMA flowchart. Excluded full-text studies and the reason for exclusion is presented in the [Additional information](https://orca.cardiff.ac.uk/id/eprint/159177/).

### 5.4 Data extraction

All demographic data was extracted directly into tables by one reviewer and checked by another. The data extracted included specific details about the populations, study methods and outcomes of significance to the review question and specific objectives. The data extraction template was piloted on manuscripts for each of the included study designs.

### 5.5 Quality appraisal

The methodological quality of all the research studies were assessed by one reviewer (and judgements verified by a second reviewer) using the JBI critical appraisal checklists specific to each research study design (https://jbi.global/critical-appraisal-tools) When a study met a criterion a score of one were given. Where a particular point was regarded as “unclear”, it was given a score of zero. Where a particular point was regarded as “not applicable” this point was taken off the total score. Overall critical appraisal scores are presented in the Additional material.

### 5.6 Synthesis

The data was reported narratively as a series of thematic summaries (Thomas et al. 2017).

### 5.7 Assessment of body of evidence

The strength of findings from the thematic summaries of RCTs (n=1) and observational studies (n=6) that provided statistical comparisons were assessed using the Grading of Recommendations, Assessment, Development and Evaluation (GRADE) approach (Guyatt et al. 2008). Due to heterogeneity of the different participant groups, and interventions outcome data was only available for results that arose from single studies and guidance was followed on undertaking the GRADE for data of this type (Ryan & Hill 2016). The resulting GRADE evidence profiles are presented in the additional information.

## EVIDENCE

### 6.1 Study selection flow chart

The PRISMA flow chart (Page et al. 2021) for the review Is displayed in Figure 1 below.

**Figure 1:**
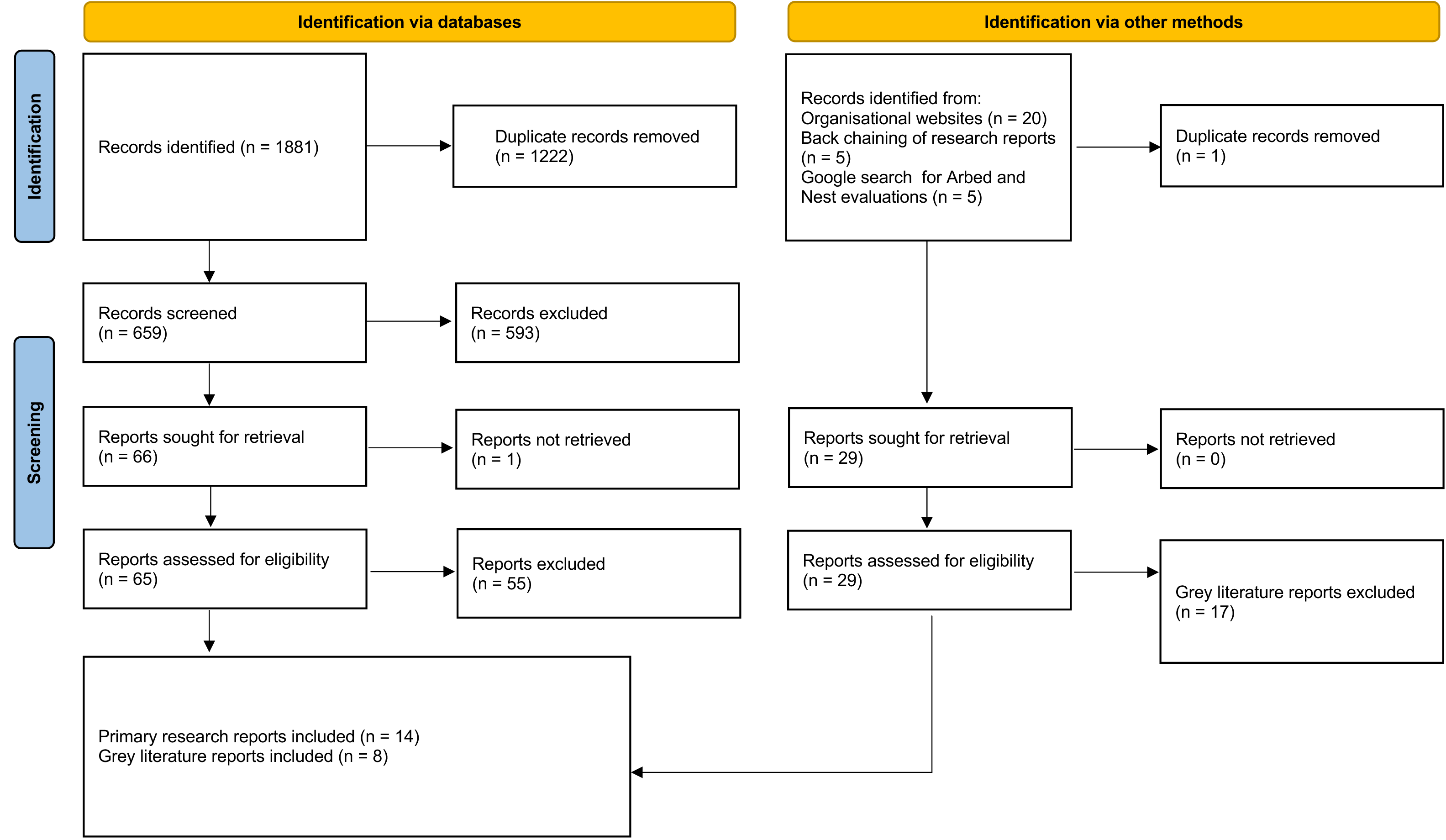
PRISMA 2020 flow diagram

### 6.2 Data extraction tables

The data extraction for the prospective and retrospective studies is displayed in Table 2 below.

**Table 2:** Summary of included evaluations.

| Citation (Country)<br>Aim<br>Url/doi | Study details | Participants & setting<br>Interventions | Key findings |
| --- | --- | --- | --- |
| Charlier et al. 2019<br>(France)<br>To assess the effectiveness of social energy subsidies and social housing to reduce fuel poverty.<br>https://doi.org/10.1080/00036846.2019.1613501 | **Study design**<br>Case control<br>**Data collection methods**<br>Questionnaire (conducted by face-to-face interview about energy consumption expenditures, attitudes, and an energy performance diagnosis of the housing)<br>**Dates of data collection**<br>2013<br>**Outcome/s of interest**<br>Energy consumption expenditures<br>Fuel poverty indicators<br>Household behaviour<br>**Outcome measures**<br>Energy-income ratio<br>Low-Income-High-Costs indicator<br>Average annual indoor temperature<br>Attitudes towards energy consumption<br>Restricted heating consumption<br>**Quality rating**<br>5 out of 10<br>JBI critical appraisal checklist for case control studies | **Participants and sample size**<br>Treatment group 1: Household in receipt of social housing without social energy subsidies (n=422)<br>Treatment group 2: Household in receipt of social energy subsidies (some of which also lived in social housing) (n=81)<br>Control: Households who live in private housing, not benefiting from social energy subsidy(n=552)<br>**Setting**<br>Rural / Urban<br>(Includes Paris and three climate areas. France is divided into three climate areas: H1 is the coldest and H3 the warmest)<br>**Type of intervention**<br>Financial: social energy subsidy<br>Energy subsidy voucher of €48 to €227 a year, depending on the household income and composition. Households will use this voucher to help pay their energy bill.<br>Energy efficient social housing | **Primary findings**<br>Results show that providing energy-efficient housing (**social housing**) to low-income households is a more efficient means of reducing fuel poverty than providing them financial support to pay their energy bills (**social energy subsidies**)<br>*Energy expenditures*<br>Living in **social housing without energy subsidies** led to a 5.4% decrease in fuel poverty with the 10% energy-income ratio approach and a 9.1% decrease with the low-income-high-cost indicator.<br>Results indicate that receiving a **social energy subsidy** does not prevent a household from being fuel poor. Benefiting from social energy subsidies has no effect on fuel poverty indicators (Energy-income ratio; Low-Income-High-Costs, even though a large part of the sample (71.6% of tenants in this treatment group) also live in social housing.<br>*Behavioral factors*<br>Households living in **social housing without energy subsidies** do not restrict their heating consumption more. On the contrary, the indoor temperature is slightly higher in social housing, by 0.29 Celsius degrees compared to controls. |
| Eadson & Leather 2017<br>(England)<br>To understand the self-assessed quantitative impacts of Fuel Poverty and Health Booster Fund | **Study design**<br>Pre-test / Post-test (no control group)<br>**Data collection methods**<br>Questionnaire (pre and 6-18 months post intervention conducted by interview)<br>**Dates of data collection** | **Participants and sample size**<br>Households where people live with health conditions that have been linked to cold homes, and people with low incomes.<br>Pre-I (n=893) / Post-I (n=193)<br>**Setting** | **Primary findings**<br>*Keeping home warm*<br>Respondents (n=193) found it easier to heat their home after receiving support)<br>Very easy (63% post-I; 5% pre-I) Fairly easy (28% post I, 6% pre-I)<br>Fairly difficult (5% post-I; 24% pre-I)<br>Very difficult (2% post-I, 3% pre-I) |
| https://www.shu.ac.uk/centre-regional-economic-social-research/publications/fuel-poverty-health-booster-fund-evaluation | Fuel Poverty and Health Booster Fund was an initiative under the 2010-2015 coalition government. No specific dates for data collection mentioned.<br>**Outcome/s of interest**<br>Keeping home warm<br>Keeping up with energy bills<br>**Outcome measures**<br>Single item questions<br>-Over the winter, how easy or difficult has it been to keep your home warm when the heating is on?<br>-How well are you and your household keeping up with your energy bills at the moment?<br>**Quality rating**<br>4 out of 9<br>JBI critical appraisal for quasi-experimental studies | Rural / Urban<br>9 local authorities received the Health Booster fund out of which analysis was commissioned for 7:<br>-Amber Valley Borough Council<br>-Derby City Council<br>-Derbyshire County Council<br>-Dudley Metropolitan Borough Council<br>-Durham County Council<br>-East Riding of Yorkshire Council<br>-Wigan Council<br>**Type of intervention**<br>Home energy advice, referral for support and/or insulation measures or home improvements: Fuel Poverty and Health Booster Fund<br>*Physical Measures*<br>Boiler (78%), central heating (18%), insulation measures (13%), loft insulation (10%), solid wall insulation (2%), double glazing (1%), cavity wall insulation (1%)<br>*Advice, support and referrals*<br>Energy saving advice (56%), use of heating system (54%), benefit entitlement (32%), switching support (30%), referral for other health intervention (18%), referral to fire service (13%), hazard checks (7%), income maximization or saving money advice (4%), debt advice (3%), advice on housing options (1%), referral for flu jab (1%) | *Keeping up with energy bills*<br>Respondents (n=193) were able to keep up with their bills better.<br>Managed well (25% post-I; 6% pre-I)<br>Managed quite well (38% pre-I; 20% post-I)<br>Get by alright (30% post-I; 38% pre-I)<br>Difficulties keeping up with bills 5% post-I; 24% pre-I)<br>-2% had severe difficulties keeping up with bills (2% post-I, 11% pre-I)<br>**Additional findings**<br>*Health and wellbeing*<br>There was a general movement towards higher scores in the post-intervention sample with regard to quality of life. |
| Grey et al. 2017 (Wales)<br>To examine the relationship between energy efficiency investments to homes in low-income areas and mental and physical health of residents, as well as a number of | **Study design**<br>Quasi-experimental<br>**Data collection methods**<br>Questionnaire (conducted before and after installation of energy efficiency measures)<br>**Dates of data collection** | **Participants and sample size**<br>Any adult resident living in the selected intervention and control areas<br>IG (n=363) / CG (n=418)<br>**Setting**<br>Urban/rural low income areas across Wales (areas were selected based on proxies of fuel | **Primary findings**<br>*Thermal satisfaction (mean±SD)*<br>IG: Baseline 3.26±1.28 / FU 4.04±1.06<br>CG: Baseline 3.61 0±1.26 / FU 3.82±1.20<br>Respondents in the intervention group were significantly more likely to report increased thermal satisfaction compared to those in the control group (OR = 3.83, 95% CI 2.40 to 5.90, Cohen's d = 0.46) |
| psychosocial outcomes likely to be part of the complex relationship between energy efficiency measures and health outcomes<br>https://doi.org/10.1186%2Fs12889-017-4075-4 | Baseline: Nov 2013 to March 2014 and Nov 2014 to Jan 2015<br>Follow-up: Nov 2014 to Jan 2015 and Nov 2015 to Dec 2015 (between 1 and 10 months post intervention)<br>**Outcome/s of interest**<br>Financial difficulties<br>Financial stress<br>Thermal satisfaction<br>Fuel poverty<br>**Outcome measures**<br>Financial difficulties scale – 4 point scale measuring how often respondents had difficulties meeting the cost of the four house related expenses of rent or mortgage payments, repairs measures or maintenance of home, fuel bills and credit card payments<br>Financial stress – 5-point scale<br>Thermal satisfaction – 5-point scale<br>Fuel poverty – single question regarding putting up with feeling cold to save heating costs.<br>**Quality rating**<br>8 out of 9<br>JBI critical appraisal checklist for quasi-experimental studies | poverty, including area deprivation, mixed tenure, and a high proportion of hard-to-heat, hard-to-treat homes)<br>**Type of intervention**<br>Energy efficiency scheme: Arbed<br>- External wall insulation (261/364, 71.7%)<br>- Full central heating (138/354, 39%)<br>- Voltage optimizer (159/358, 44.4%)<br>- Heating control (101/353, 28.6%)<br>- Connection of off-gas communities to the mains gas network (49/353, 13.95) | *Fuel poverty (mean±SD)*<br>IG: Baseline 0.63±0.48 / FU 0.45±0.50<br>CG: Baseline 0.57±0.50 / FU 0.46±0.50<br>The number of respondents reporting putting up with feeling cold to save heating costs significantly decreased for the intervention group compared to the control group (OR 0.49, CI 0.25 to 0.94, Cohen's d = 0.15).<br>*Financial difficulties (mean±SD)*<br>IG: Baseline 1.93±0.90 / FU 1.67±0.73<br>CG: Baseline 1.74±0.75 / FU 1.65±0.73<br>Those in the intervention group reported significantly fewer financial difficulties compared to those in the control group (B=-0.15, 95% CI -0.25 to -0.05, Cohen's d = 0.20).<br>*Financial stress (mean±SD)*<br>IG: Baseline 2.96±1.40 / FU 2.60±1.35<br>CG: Baseline 2.81±1.38 / FU 2.58±1.36<br>No statistical analysis reported only Cohen's d = 0.10 |
| Heyman et al. 2011 (England)<br>To assess the potential health, and also personal, social and economic benefits of energy efficiency measures<br>https://doi.org/10.1080/02673037.2010.512787 | **Study design:**<br>Randomised control trial (with partial crossover)<br>**Data collection methods**<br>Questionnaire (conducted by interview about satisfaction with energy efficiency)<br>**Dates of data collection**<br>2000 – 2004 (4-year period)<br>**Outcome/s of interest** | **Participants and sample size**<br>Households living in full or marginal fuel poverty; data was collected for four years, with year one and two being baseline measurements. The IG received home improvements in year 3 but not the control to allow for comparison, while the CG received measures in year 4<br>Year one: IG (n=129) / CG (n=108)<br>Year two: IG (n=114) / CG (n=92)<br>Year three: IG (n=99) / CG (n=83) | **Primary findings:**<br>*Mean SAP Energy efficiency rating (Year 3)*<br>IG: (n=96, 61.1±13.5) / CG: (n=82, 48.5±11.6 (p<0.001)<br>*Difference between living room and external temperature (7am to 10am)*<br>IG: 13.8°C / CG: 13.0 °C; p=0.10<br>*Difference between living room and external temperature (6pm to 11pm)*<br>IG: 14.5°C / CG: 13.1°C; p=0.03 |
|  | Energy efficiency rating<br>Room temperature<br>Average fuel expenditure<br>Satisfaction with home warmth<br>**Outcome measures**<br>SAP rating system<br>Fuel costs calculated from meter readings/fuel bills.<br>Satisfaction with home heating 8-item scale.<br>**Quality rating**<br>6 out of 10<br>JBI critical appraisal checklist for randomised controlled trials. | Year four: IG (n=70) / CG (n=70)<br>**Setting**<br>Rural / Urban<br>Relatively poor area of Tyne and Wear in North East England<br>**Type of intervention**<br>Energy efficiency scheme: retrofitting (worth an average of £727 (range £0-£3,335)<br>- Loft insulation (54%) /<br>- Cavity wall insulation (53%)<br>- Draught exclusion (29%)<br>- Heating controls (20%)<br>- Central heating (13%)<br>and other measures as required. | *Difference between bedroom and external temperature (10pm to 9am)*<br>IG: 12.9°C / CG: 12.3 °C; p=0.26<br>*Average fuel expenditure*<br>IG n=99, £596 / CG n=83, £567, p=0.408<br>*Improvement in satisfaction with home warmth (comparing years two and three),*<br>IG :1.18 points / CG 0.64; (p=0.02)<br>**Additional findings**<br>% reporting problems with heating system<br>IG: 11.2%, n=10 / CG: 22.4%, n=17; p=0.05 |
| McGinley et al. 2022 (Ireland)<br>To develop an integrated framework of key performance indicators which are to be used to demonstrate the wider benefits of retrofitting.<br>https://doi.org/10.3390/en15010334 | **Study design**<br>Case study<br>**Data collection methods**<br>Building characteristic survey<br>Questionnaire<br>Temperature, relative humidity, and electricity consumption data-logging instrumentation<br>Monthly readings of electricity and oil usage<br>**Dates of data collection**<br>Pre and post retro survey.<br>December 2015 to February 2018<br>**Outcome/s of interest**<br>Thermal comfort (household's thermal satisfaction within their homes).<br>Expected fuel poverty alleviations.<br>Household self-reported energy use.<br>**Outcome measures**<br>Quantitative temperature data<br>Fuel poverty status | **Participants and sample size**<br>Adults in each household (n=5) where retrofit had taken<br>**Setting**<br>Rural (detached rural dwellings)<br>Case A: a solid masonry wall house constructed in the 1960s (94 m<sup>2</sup> heated floor area)<br>Case B: two-storey detached house (118 m<sup>2</sup> heated floor area)<br>Case C: bungalow home constructed in the 1950s (107 m<sup>2</sup> heated floor area)<br>Case D: bungalow constructed in the late 1970s (72 m<sup>2</sup> heated floor area)<br>Case E: bungalow constructed in the 1960s (71 m<sup>2</sup> heated floor area)<br>**Type of intervention**<br>Energy efficiency scheme: retrofitting<br>Case A: external insulation, double glazing, heating system<br>Case B: heating system<br>Case C: loft insulation, heating system<br>Case D: space and water heating system<br>Case E: heating system | **Primary findings**<br>*Reduction in energy usage*<br>Case A: 65% reduction in primary energy usage, and 71% reduction in secondary energy usage<br>Case B: 66% reduction in primary energy usage, and 70% reduction in secondary energy usage<br>Case C: 1% reduction in primary energy usage, and 3% reduction in secondary energy usage<br>Case D: 21% reduction in primary energy usage, and 20% reduction in secondary energy usage<br>Case E: 43% reduction in primary energy usage, and 59% reduction in secondary energy usage<br>*Fuel poverty status*<br>It is expected that only Case A and B would not be in fuel poverty over a 30-year period following retrofit measure installation.<br>*Thermal comfort*<br>Case A: 19% increase in indoor temperature<br>Case B: 4% decrease in indoor temperature<br>Case C: 6% increase in indoor temperature<br>Case D: 1% increase in indoor temperature<br>Case E: 7% increase in indoor temperature |
|  | Reduction in primary or secondary energy usage (annual heating energy usage or electricity and oil energy usage levels depending on what is used primarily)<br>**Quality rating**<br>7 out of 8<br>JBI critical appraisal checklist for case reports |  |  |
| Miller et al. 2022 (Wales)<br>To undertake a process and impact evaluation of the Arbed 3 programme<br>https://www.gov.wales/sites/default/files/statistics-and-research/2022-02/evaluation-of-arbed-3.pdf | **Study design**<br>Quantitative descriptive survey as part of a wider mixed method evaluation<br>**Data collection methods**<br>Questionnaire<br>**Dates of data collection:**<br>Interim evaluation (Apr 2020 – Mar 2021)<br>Final impact evaluation (Jan - July 2021)<br>**Outcome/s of interest**<br>Average reduction in energy costs<br>EPC ratings<br>**Outcome measures**<br>Uses estimates of fuel poverty<br>Proportion of income spent on energy: answer categories (10%, between 10 and 20% and more than 20%)<br>Improvement in EPC ratings<br>**Quality rating**<br>5 out of 8<br>JBI critical appraisal checklist for analytical cross-sectional studies | **Participants and sample size**<br>Households experiencing fuel poverty: characterized by having an EPC rating of E, F or G EPC rating (subsequently revised to permit a proportion of D rated households) (n=506)<br>**Setting**<br>Urban and rural areas of Wales (deprived areas)<br>West Wales and Valley, East Wales with the majority of households supported from Rhondda Cynon Taf (RCT), Flintshire, and Blaenau Gwent.<br>**Type of intervention**<br>Energy efficiency scheme: Arbed | **Primary findings**<br>*Self-reported reduction in energy costs*<br>44% felt that their energy bills had been reduced. 30 per cent of survey respondents stated that they spent more than 20 per cent of their income on energy bills pre-installation (indicative of severe fuel poverty), this reduced to 5 per cent post-installation, demonstrating the likely impact the measures had on increasing monthly disposable income.<br>Following the installation of measures, 61 per cent of householders claimed they now spend less than 10 per cent of their household income on energy bills every month, compared to 31 percent pre-installation.<br>*Energy efficiency*<br>Of the 2,546 properties receiving measures, from which all experienced a minimum one band uplift, 70 per cent experienced an uplift of one EPC band, 24 per cent were raised two bands, five per cent three bands and 1 percent above 3 bands. |
| Papada et al. 2021 (Greece) | **Study design**<br>Pre-test / Post-test (no control group)<br>**Data collection methods** | **Participants and sample size**<br>Energy vulnerable households (n=150)<br>**Setting** | **Primary findings**<br>*Energy-related behaviour* |
| To contribute to the body of knowledge of energy vulnerability in Greek mountainous areas through the operation of a living lab<br>https://doi.org/10.3390/en14061525 | Questionnaire<br>**Dates of data collection**<br>3 independent rounds:<br>(1) March – August 2019<br>(2) November 2019 – May 2020<br>(3) July – December 2020<br>**Outcome/s**<br>Energy-related behaviour<br>- behaviour patterns / adoption of measures and advice<br>Energy costs for heating and electricity<br>Energy vulnerability<br>**Outcome measures**<br>Thermal discomfort or inability to keep home adequately warm.<br>Housing condition, including moisture / mold problems<br>Arrears in energy bills<br>% increase in burner efficiency (energy-efficiency ratio)<br>Energy savings (calculated on the basis of each household's specific energy consumption, as estimated by the energy advisors (kWh/Year)<br>Reduction in energy costs (Euros/year)<br>**Quality rating**<br>4 out of 9<br>JBI critical appraisal checklist for quasi-experimental studies | Remote - Metsovo: mountainous town of Greece<br>**Type of intervention**<br>Living lab - Installing monitoring equipment, inspecting heating systems and specific household energy advice<br>- Group a: Energy advisors visits and advice and monitoring equipment installed<br>- Group b: Energy advisors visits, and advice and no monitoring equipment installed<br>Further advice was also provide delivered at three energy cafes | 85.5% of households stated that the monitoring equipment motivated them toward taking certain energy-efficiency decisions:<br>- maintenance/change of the heating system (15%)<br>- change of analogue to digital thermostats (14%)<br>- insulation measures (13%)<br>- purchase of some energy-efficient appliance (12%)<br>- change of light bulbs (9%)<br>- change of habits/reduction in consumption (7%)<br>- change of time-of-use of home appliances (6%)<br>- purchase of dehumidifiers (5%)<br>Households in group A were significantly more likely to plan to apply energy efficiency measures compared to those in group B (80% vs. 23%, p=0.000)<br>*Energy savings and reduction in heating costs (due to maintenance of central heating systems in 12 households)*<br>Significant increase in the burners' energy-efficiency ratio<br>Energy savings 11,532kWh/Year<br>Reduction in energy costs 1286 Euros<br>*Thermal comfort*<br>40% of households reported an improvement in their quality of life due mainly to an improvement in the level, of thermal comfort at home (42%), by facing less moisture problems (26%) and by reducing energy costs (23%)<br>**Additional findings**<br>*Improvement of living conditions*<br>Households in group A were significantly more likely to report an improvement in their living conditions compared to those in group B (58% vs. 25%, p=0.000) |
| Poortinga et al. 2017 (Wales)<br>To examine the impacts of the intervention programme on internal hydrothermal conditions and energy use in low-income households | **Study design**<br>Quasi-experimental<br>**Data collection methods**<br>Long-term monitoring of the indoor environment in two subsequent heating (winter) seasons<br>**Dates of data collection**<br>November 2014 to April 2015 | **Participants and sample size**<br>Low income households<br>Baseline: IG (n=50) / CG (n=49)<br>Follow-up: IG (n=48) / CG (n=40)<br>**Setting**<br>Low income areas urban/rural areas in Wales | **Primary findings**<br>*Overall indoor air temperature at follow-up (Mean±SD)*<br>IG: 18.95±2.37 °C / CG: 17.38±2.66 °C<br>Relative change (K): 0.84; 95% CI [0.64, 1.04]; p<0.001<br>*Daily average living room temperature at follow-up*<br>IG: 19.33±2.68 °C / CG: 18.26±2.77 °C<br>Relative change (K): 1.01; 95% CI [0.78, 1.23]; p<0.001<br>*Daily average bedroom temperature at follow-up* |
| https://doi.org/10.1080/09613218.2017.1314641 | **Outcome/s of interest**<br>Indoor air temperature at different times of day and in different rooms<br>Substandard internal conditions (recording the time each day the indoor air temperature dropped below 16 or 18°C, and indoor relative humidity was above 60%)<br>Gas usage<br>**Outcome measures**<br>Temperature data recorded at different times of the day using Tinytag Ultra 2 data loggers.<br>Average daily gas usage using meter readings.<br>**Quality rating**<br>8 out of 9<br>JBI critical appraisal checklist for quasi-experimental studies | Brynamman (Carmarthenshire), Caerau (Cardiff), Llay (Wrexham), Hollybush (Caerphilly), and Pennydarren(Merthyr Tydfil)<br>**Type of intervention**<br>Energy efficiency scheme: Arbed<br>The study was conducted in five low income areas where Arbed was scheduled to take place (IG), and five comparable control areas where no such investments were planned during the duration of the study (CG)<br>Internal conditions were monitored for a minimum of 28 consecutive days before and after the installation of energy-efficiency measures, and compared those with internal conditions of households that did not receive such measures.<br>*Two measures (n=32/48)*<br>External insulation and heating system (n=19)<br>Gas mains network and new heating system (=13)<br>*Three measures (n=16/48)*<br>External insulation, heating system, new windows and doors (n=9)<br>External insulation, gas mains network, heating system (n=7) | IG: 18.86±2.91 °C / CG: 16.64±3.22 °C<br>Relative change (K): 1.28; 95% CI [1.04, 1.52]; p<0.001<br>*Daily average kitchen temperature at follow-up*<br>IG: 18.68±2.69 °C / CG: 17.25±3.39 °C<br>Relative change (K): 0.24; 95% CI [-0.01, 0.48]; p=0.060<br>*Substandard internal conditions at follow-up*<br>Percentage distribution of indoor air temperature<br>IG: 11.0 / CG: 30.4 < 16°C<br>IG: 18.5% / CG: 26.6% 16°C-18°C<br>IG: 68.5 / C: 42.9% 18 °C -24 °C<br>IG: 1.2 / CG: 0.1 >24 °C<br>$\chi^2(3) = 1.761$, p = 0.623<br>*Length of substandard internal conditions*<br>Relative change (hour < 16°C): 0.20; 95% CI [-0.48, 0.88]; p=0.567<br>Relative change (hour < 18°C): 0.27; 95% CI [-0.49, 1.03]; p=0.483<br>*Cumulative substandard conditions*<br>Relative change (K hour < 16°C for hours): -4.20; 95% CI [-6.64, -1.76]; p< 0.001<br>Relative change (K hour < 18°C): -3.62; 95% CI [-6.95, -0.30]; p=0.003<br>*Change in average daily gas use*<br>IG (n=26): Baseline: 3.88 m<sup>3</sup>; Follow-up: 2.45 m<sup>3</sup>; Change: 36.9%<br>Repeated measures ANOVA: p=0.000, (Cohen's d = 1.41)<br>**Additional Findings:**<br>*Indoor air temperature change depending on measures installed*<br>Relative change (external insulation): 1.12; 95% CI [0.69, 1.55]; p< 0.001<br>Relative change (windows and doors): -0.02; 95% CI [-0.39, 0.35]; p=0.924<br>Relative change (heating system): -0.19; 95% CI [-0.69, 0.31]; p< 0.463<br>Relative change (gas network): 0.69; 95% CI [0.29-1.09]; p< 0.001 |
|  |  |  | *Indoor air temperature change depending on building construction*<br>Relative change (cavity wall): -0.17; 95% CI [-0.58, 0.25]; p=0.430<br>Relative change (solid wall): 0.74 ; 95% CI [0.51, 0.96]; p<0.001<br>Relative change (British steel framed): 1.54; 95% CI [1.26–1.83] ; p<0.001 |
| Sharpe et al. 2020 (England)<br>To assess the impact of an intervention to install a new first time central heating system in order to reduce fuel poverty on household satisfaction with indoor temperatures/ environment, ability to pay bills and mental wellbeing<br>https://doi.org/10.1177/1420326X20975468 | **Study design**<br>Post-test - with control group<br>**Data collection methods**<br>Questionnaire<br>**Dates of data collection**<br>July and August 2019<br>**Outcome/s of interest**<br>Ability to pay bills<br>Avoiding heating due to costs<br>**Outcome measures**<br>Single item questions<br>- Compared to this time last year have you had problems with paying your bills? (“Worsened a lot” to “Worsened a little”; “Not changed”; “Improved a little” to “Improved a lot”)<br>- Do you avoid turning on the heating because of cost? (Yes or No)<br>**Quality rating**<br>4 out of 9<br>JBI critical appraisal checklist for cohort studies (prospective) | **Participants and sample size**<br>Lower income population who had applied for a local authority declaration to receive support from the ECO Flex programme<br>- IG (n=71)<br>- CG (Waiting list control) (n=83)<br>**Setting**<br>Rural: Cornwall<br>**Type of intervention**<br>Energy efficiency measure: central heating<br>Qualifying households received support and funding towards a new first time heating system under the new ECO Flex programme<br>The funding allowed the installation of a new central heating system in homes reliant on a single source of heating in one room or those using electric heating | **Primary findings**<br>*Ability to pay bills*<br>CG: worsened a lot to a little (30.77%); No change (41.03%); Improved (**21.21%**)<br>IG: worsened a lot to a little (8.96%); No change (40.30%); Improved (**50.75%**)<br>Chi squared test p<0.01<br>*Avoiding heating due to costs*<br>CG: No (16.46%); Yes (**83.54%**)<br>IG: No (41.79%); Yes (**58.21%**)<br>Chi squared test p<0.01<br>**Additional findings**<br>*Mental wellbeing (Mean+SD)*<br>CG: 21.50+0.65 (n=69) / IG: 21.48+0.55 (n=58), p=0.99<br>*Percentage with low mental wellbeing*<br>In households with a new heating system there were fewer participants with a low mental wellbeing (4.23% when compared to the control (18.07%) (p<0.01) |
| Sherriff et al. 2020 (Scotland)<br>To evaluate the Gluasad Còmhla (Moving Together) project | **Study design**<br>Quantitative descriptive survey as part of a wider mixed methods study<br>**Data collection methods**<br>Questionnaire (postal or online) | **Participants and sample size**<br>People in fuel poverty whose health is compromised by living in a cold or hard to heat home and have long-term conditions (n=27/198 response rate 14%) | **Primary findings**<br>*Use of heating system*<br>Much more often (n=3, 11%); Slightly more often (n=0); About the same (n=10, 37%); Slightly less often (n=6, 22%); Much less often (n=1, 4%) |
| https://usir.salford.ac.uk/id/eprint/57055/1/Sherriff%202020%20Moving%20Together%20Final%20Rep | **Dates of data collection**<br>January 2020<br>**Outcome/s of interest**<br>Use of heating system<br>Temperature on a cold day<br>Cost of running the heating system<br>Effects of temperature on indoor activities<br>Money available after bills<br>**Outcome measures**<br>Single item questions (compared to before)<br>do you now use your heating system...?<br>- how much does your heating system cost to run...?<br>- does the temperature of your home affect what you do at home?<br>- how much money do you have available to spend once you have paid your bills?<br>**Quality rating**<br>5 out of 8<br>JBI critical appraisal checklist for analytical cross sectional studies | **Setting**<br>Rural and remote areas: Outer Hebrides<br>**Type of intervention**<br>Home energy advice, referral for support and/or insulation measures or home improvements:<br>Glusad Còmhlà (Moving Together) built on the existing practice of TIG. TIG is a Community Benefit Society supporting people to access homes and to help to make them comfortable and affordable, promote independent living and encourage businesses and communities to be energy-efficient.<br>Home energy visit and/or advice (n=127, 64%)<br>Referral onto other assistance (n=69, 35%)<br>Help with bills, including switching (n=45, 23%); Grants and benefits (n=32, 16%)<br>Insulation (n=26, 13%)<br>Other (n=9, 5%);<br>Other equipment or technology (n=6, 3%) | *Temperature on a cold day*<br>Much warmer (n=7, 26%); Slightly warmer (n=8, 30%); About the same temperature (n=4, 15%); Slightly colder (n=0); Much colder (n=1, 4%)<br>*Cost of running the heating system*<br>Much more (n=1, 4%); Slightly more (n=2, 7%); About the same (n=9, 33%); Slightly less (n=7, 26%); Much less (n=2, 7%)<br>*Effects of temperature on indoor activities*<br>Much more often (n=0); Slightly more often (n=4, 15%); About the same (n=15, 56%); Slightly less often (n=0); Much less often (n=1, 4%)<br>*Money available after bills*<br>Much more (n=2, 7%); Slightly more (n=6, 22%); About the same (n=8, 30%); Slightly less (n=3, 11%); Much less (n=2, 7%)<br>**Additional findings**<br>*Overall Health*<br>Much better (n=1, 4%); Slightly better (n=4, 15%); About the same (n=11, 41%); Slightly worse (n=2, 7%); Much worse (n=3, 11%) |
| Shortt & Rugkasa 2007<br>(Northern Ireland)<br>To report findings of an evaluation of a fuel poverty programme in the Armagh and Dungannon Health Action Zone in Northern Ireland<br>https://doi.org/10.1016/j.healthplace.2005.10.004 | **Study design**<br>Pre-test / Post-test with a control group as part of a wider mixed methods study<br>**Data collection methods**<br>Questionnaire covering a time period both pre and post intervention (n=12)<br>**Dates of data collection**<br>Pre intervention questionnaire<br>July 2000 to August 2002<br>Post intervention questionnaire at 1 year follow up<br>May 2003 – January 2004<br>**Outcome/s of interest** | **Participants and sample size**<br>Households in fuel poverty<br>As some households received a heating system (total intervention) and some did not, those who did not were seen as the CG (n=46) and those who did the IG (n=54) (numbers reflect those who replied in the post intervention survey)<br>**Setting**<br>Rural: Armagh and Dungannon Health Action Zone<br>**Type of intervention**<br>Energy efficiency measure: central heating | **Primary findings**<br>*Results from the temperature data loggers (n=12)*<br>There was relatively little change in the average temperatures pre and post intervention<br>Of the 56 total intervention households, 26 restricted heating to 1-2 rooms prior to the intervention. After heating systems were installed, this has reduced to 1 household<br>*Fuel Costs*<br>Drop in fuel costs was most significant for total intervention households whose average household fuel costs fell significantly from £1113 per annum to £751.56 per annum (p<0.001)<br>*Households were claiming some form of benefit* |
|  | Temperature of the home<br>Uptake of benefits<br>**Outcome measures**<br>Temperature data loggers<br>Estimate how much they spent on fuel (for both heating and other use) for each type of fuel consumed<br>**Quality rating**<br>4 out of 9<br>JBI critical appraisal checklist for quasi-experimental studies |  | CG: Pre-intervention (2%); post-intervention (58%), p=0.001<br>IG: Pre-intervention (98%)<br>*Mean number of benefits per house*<br>CG: Pre-intervention (0.02); post-intervention (0.71), p=0.001<br>IG: Pre-intervention (1.78)<br>**Additional findings**<br>*Health*<br>For total intervention households there was a significant decrease in both the numbers of householders reporting arthritis/rheumatism (p<0.05) and the numbers reporting 'other' form of illness (p<0.05) |
| Wade et al. 2019<br>(Scotland)<br>To test the Energycarer approach to tackling rural fuel poverty in two rural areas<br>https://www.gov.scot/publications/evaluation-hes-homecare-pilot/ | **Study design**<br>Pre-test / post-test with control group as part of a wider mixed method evaluation<br>**Data collection methods**<br>Internal temperature monitoring<br>Questionnaire<br>**Dates of data collection**<br>March 2017 to 31<sup>st</sup> March 2019<br>**Outcome/s of interest**<br>Internal temperature<br>Change in thermal comfort during winter months<br>Energy related behaviour change<br>**Outcome measures**<br>Tinytag temperature monitors<br>Single item questions<br>-During the colder winter months (October to March), how would you describe the thermal comfort of your home?<br>(Ordinal scale from "Much too cool" to "Much too warm") | **Participants and sample size**<br>Rural populations with high incidence of fuel poverty who had received the HES homecare service<br>IG: (n=11) / CG: (n=3) (out of these only three from the IG were analysed)<br>Questionnaire<br>Baseline: IG: (n=17) / CG: (n=7)<br>Follow-up: IG: (n=14) / CG: (n=4)<br>**Setting**<br>Rural areas of Anandale & Eskdale (South West Scotland) and Moray East (North East Scotland)<br>**Type of intervention**<br>Home energy advice, referral for support and/or insulation measures or home improvements<br>Two Energycarers acted as case workers providing individually tailored solutions to clients with the aim of delivering affordable warmth (ranging from 3 to 12 visits).<br>Energycarers during initial visits assessed client's needs, their home, looked for insulation measures or heating improvements that could | **Primary findings**<br>*Internal temperature before and after installation*<br>Property A (Gas Boiler): 1% change<br>Property B (Draughtproofing): -1% change<br>Property C (Electric Storage Heaters): 1% change<br>No significant change was found (p>0.05), although the sample size (n=3) is too small for appropriate statistical analysis<br>*Change in thermal comfort during winter months*<br>IG: no change (n=7); substantial improvement (n=2); minor improvement (n=2); deterioration (n=1)<br>CG: no change (n=2); deterioration (n=2); minor improvement (n=1)<br>*Energy related behaviour change*<br>Some modest changes were reported in people's actions, such as using extra clothing and outdoor clothing less (responses changing from "very often" to, "often" for those in the IG) No overall change and no statistical evidence reported. |
|  | -Thinking back to last winter, how frequently did you do the following to prevent yourself being too cold at home?<br>**Quality rating**<br>5 out of 9<br>JBI critical appraisal checklist for quasi-experimental studies | be installed, assessed the householder's ability to pay for measures and checked whether clients qualified for Warmer Homes Scotland. Those who did not qualify for Warmer Homes Scotland were referred for physical works to Warmworks, Care and Repair Moray, or Care and Repair Dumfries and Galloway depending on which was most appropriate.<br>Those who took part in the internal temperature monitoring received the following heat related energy efficiency measures following Energycarer visits:<br>boiler replacement (n=5); glazing/doors (n=4); electric heating upgrade (n=2); draught proofing (n=2); hot water cylinder (n=2); loft insulation (n=1); electric shower (n=1)<br>The control group were part of the CLO programme |  |
| Welsh Government 2015 (Wales)<br>To assess the extent to which the scheme has met its objectives and further, to provide evidence of its impact and effectiveness<br>https://www.gov.wales/sites/default/files/statistics-and-research/2019-07/150310-evaluation-nest-energy-efficiency-scheme-en.pdf | **Study design**<br>Quantitative descriptive survey as part of a wider mixed method evaluation<br>**Data collection methods**<br>Questionnaires (conducted by telephone)<br>**Dates of data collection**<br>September 2014<br>**Outcome/s of interest**<br>Economic<br>Behaviour change<br>Value for money<br>**Outcome measures**<br>Perceived ability to heat homes<br>Energy bill reduction<br>Energy use<br>Annual energy saving<br>Annual rate of return<br>**Quality rating** | **Participants and sample size**<br>Home improvement beneficiaries (n= 600 / 9506 households)<br>Advice and guidance beneficiaries (n=1000 individuals)<br>**Setting**<br>Households in Wales<br>(Total number of households receiving measures n=15677 (rural n=5776; 37%))<br>**Type of intervention**<br>Energy efficiency scheme: Nest<br>Home improvement:<br>Home energy advice and guidance<br>*Total number of measures installed:*<br>Gas boilers (62.3%) / Oil (11.3%)<br>Solid fuel (3.2%) / Electric (3.0%)<br>LPG (2.6%) / Air source heat pumps (1.1%)<br>Biomass (0.4%) / Loft insulation (9.9%)<br>Cavity wall insulation (2.4%) | **Primary findings**<br>89% reported that they were better able to heat their home as a result of the installation<br>62% reported that their energy bills had been reduced as a result of the household improvements<br>35% reported that they were better able to heat their home as a result of the advice provided by Nest<br>83% stated that they were more aware of their energy use as a result of the installation they received<br>*Value for money*<br>Estimation the overall annual energy saving accruing to beneficiary households is £7.48m<br>Overall investment of £58m - equates to a benefit to cost ratio of 1.29, meaning that the Welsh economy achieves a return of £1.29 back for each £1 spent<br>Alternatively, this could be seen as having a 7.75 year payback period, or an annual rate of return of 12.9 per cent |
|  | 5 out of 8<br>JBI critical appraisal checklist for analytical cross sectional studies | Draft proofing (1.9%) / External insulation (1.7%)<br>Cylinder insulation (0.1%) | **Additional findings**<br>Almost half of beneficiaries reported improvements in their health or that of their family as a result of the home improvements |
| Willand et al. 2019<br>(Australia)<br>To quantify changes in indoor temperatures, energy consumption, energy costs and health due to building retrofits<br>https://doi.org/10.1016/j.erss.2019.02.017 | **Study design**<br>Quasi-experimental study as part of a wider mixed methods study<br>**Data collection methods**<br>Questionnaires<br>Data loggers (temperature measurements)<br>**Dates of data collection**<br>Before and after the retrofit interventions: 2014 and 2015<br>**Outcome/s of interest**<br>Energy efficiency<br>Energy consumption and affordability<br>Household temperatures<br>**Outcome measures**<br>Home energy efficiency ratings<br>Billed consumption data<br>Quantitative temperature readings<br>**Quality rating**<br>5 out of 9<br>JBI critical appraisal checklist for quasi-experimental studies | **Participants and sample size**<br>Low income, older and/or frail elderly home care recipients. (Home care services is defined as services offering domestic cleaning services, home maintenance and modification for better accessibility). 30 households recruited (one dropped out)<br>IG: (n=13) / CG: control (n=16)<br>**Setting:**<br>Urban/Rural: Victoria, Australia<br>**Type of intervention:**<br>Energy efficiency scheme: retrofits<br>- replacement of lightbulbs with LED's<br>- top-up R4 ceiling/roof insulation and draught proofing<br>- new reverse cycle air conditioner<br>CG households received retrofit at the end of the study period | **Primary findings**<br>*Energy efficiency (star ratings)*<br>IG: pre retro-fit (n=10): mean 2.7 stars<br>IG: post retro-fit (n=9) mean 3.5 stars<br>CG: (n=9) mean 2.9 stars<br>*Indoor temperatures*<br>The analysis was not able to provide evidence for statistically significant benefits of the intervention measures on various indicators of living (p=.53 and r=.21) or bedroom warmth (p=0.933, r=0.05)<br>*Mean changes in electricity consumption*<br>CG: (n=13) +15% / IG (n=16) -6%, p=0.017, r=0.44)<br>*Mean changes in gas consumption*<br>CG: (n=13) + 7% / IG (n=16) +5% (r=0.08, p=0.742) |

### 6.3 Information available on request

The protocol is available on request. Search strategies, list of excluded studies, critical appraisal, and GRADE evidence profiles are available here: [Additional information](https://orca.cardiff.ac.uk/id/eprint/159177/)

## 6.5 Acknowledgements

The authors would like to thank James Burgess, Brendan Collins, Tom Smithson, Rebecca Masters, and Debs Smith for their expertise, time and contributions during stakeholder meetings in guiding the focus of the review and interpretation of findings.

## 7. ABOUT THE HEALTH AND CARE RESEARCH WALES EVIDENCE CENTRE

The Health and Care Research Wales Evidence Centre integrates with worldwide efforts to synthesise and mobilise knowledge from research.

We operate with a core team as part of [Health and Care Research Wales](https://healthandcareresearchwales.org/about-research-community/wales-covid-19-evidence-centre), Welsh Government, and are led by [Professor Adrian Edwards of Cardiff University](https://profiles.cardiff.ac.uk/staff/edwardsag).

The core team of the centre works closely with collaborating partners in the [Bangor Institute for Health and Medical Research (BIHMR)](https://www.bangor.ac.uk/shs/research), Bangor University, which includes the [Centre for Health Economics and Medicines Evaluation (CHEME)](https://cheme.bangor.ac.uk/) working in collaboration with Health and Care Economics Cymru, [Health Technology Wales](https://healthtechnology.wales/), [Public Health Wales Evidence Service](https://phw.nhs.wales/services-and-teams/observatory/evidence/), [Population Data Science, Swansea University](https://popdatasci.swan.ac.uk/about-us/) using [SAIL Databank](https://saildatabank.com/), the [Wales Centre for Evidence Based Care (WCEBC)](https://www.cardiff.ac.uk/healthcare-sciences/about-us/engagement/the-wales-center-for-evidence-based-care-wcebc), the [Specialist Unit for Review Evidence (SURE)](https://www.cardiff.ac.uk/specialist-unit-for-review-evidence) and [CASCADE](https://cascadewales.org/), Cardiff University.

**Director:** Professor Adrian Edwards

**Website:** www.researchwalesevidencecentre.co.uk

## APPENDIX

## APPENDIX 1: Grey literature resources searched

ADR UK - https://www.adruk.org/

Campbell collaboration - https://www.campbellcollaboration.org/

Centre for Sustainable Energy - www.cse.org.uk/

Citizens Advice (Scotland) - https://www.cas.org.uk/

Citizens Advice (Wales) - https://www.citizensadvice.org.uk/wales/

Department for Business, Energy & Industrial Strategy - https://www.gov.uk/government/organisations/department-for-business-energy-and-industrial-strategy

Department of Energy and Climate Change - https://www.gov.uk/government/organisations/department-of-energy-climate-change

End Fuel Poverty Coalition - www.endfuelpoverty.org.uk/

Energy Action Scotland - https://www.eas.org.uk/eas-publications

Gov.uk - www.gov.uk

https://www.scotphn.net/groups/scottish-directors-of-public-health/introduction/

Institute of Health Equity - https://www.instituteofhealthequity.org/home

Joseph Rowntree Foundation - https://www.jrf.org.uk/

Local Government Association - https://www.local.gov.uk/

National Energy Action - https://www.nea.org.uk/

Northern Ireland Public Health Agency - https://www.publichealth.hscni.net/

Office for Health Improvement and Disparities - https://www.gov.uk/government/organisations/office-for-health-improvement-and-disparities

Ofgem - www.ofgem.gov.uk/

Policy Exchange - https://policyexchange.org.uk/

Public Health Scotland - https://publichealthscotland.scot/

Public Health Scotland - https://www.scotphn.net/

Public Health Wales - https://phw.nhs.wales/

Scottish Directors of Public Health Scottish Government - https://www.gov.scot/

Shelter Scotland - https://scotland.shelter.org.uk/

The Bevan Foundation - https://www.bevanfoundation.org/

The International Public Policy Observatory - https://covidandsociety.com/

UK Parliament - https://www.parliament.uk/

Wales Centre for Public Policy - https://www.wcpp.org.uk/publications/

Welsh Government - https://gov.wales/

Welsh Parliament - https://senedd.wales/

